# Multi-layered Diagnostic Protocol Improves Postsurgical Outcomes in Children with Drug-resistant Epilepsy And Focal Cortical Dysplasia Type 1

**DOI:** 10.1101/2024.09.24.24314277

**Authors:** Barbora Splitkova, Katerina Mackova, Miroslav Koblizek, Zuzana Holubova, Martin Kyncl, Katerina Bukacova, Alice Maulisova, Barbora Straka, Martin Kudr, Matyas Ebel, Alena Jahodova, Anezka Belohlavkova, Gonzalo Alonso Ramos Rivera, Martin Hermanovsky, Petr Liby, Michal Tichy, Josef Zamecnik, Radek Janca, Pavel Krsek

## Abstract

**Objectives:** We comprehensively characterised a large paediatric cohort with histologically confirmed focal cortical dysplasia (FCD) type 1 to demonstrate the role of advanced multimodal pre-surgical evaluation and identify predictors of postsurgical outcomes.

**Methods:** This study comprised a systematic re-analysis of clinical, electrophysiological, and radiological features. The results of this re-analysis served as independent variables for subsequent statistical analyses of outcome predictors.

**Results:** All children (N = 31) had drug-resistant epilepsy with varying impacts on neurodevelopment and cognition (presurgical intelligence quotient (IQ)/developmental quotient scores: 32–106). Low presurgical IQ was associated with abnormal slow background electroencephalogram (EEG) activity and disrupted sleep architecture. Scalp EEG showed predominantly multiregional and often bilateral epileptiform activity. Advanced epilepsy magnetic resonance imaging (MRI) protocols identified FCD-specific features in 74.2% of patients (23/31), 17 of whom were initially evaluated as MRI-negative. In six out of eight MRI-negative cases, fluorodeoxyglucose positron emission tomography (FDG-PET) and subtraction ictal single-photon emission computed tomography co-registered to MRI (SISCOM) helped localise the dysplastic cortex. Sixteen patients (51.6%) underwent stereoelectroencephalography (SEEG). Twenty-eight underwent resective surgery, and three underwent hemispheral disconnection. Seizure freedom was achieved in 71.0% of patients (22/31) by the last follow-up, including seven of the eight MRI-negative patients. Anti-seizure medications (ASMs) were reduced in 21 patients, with complete withdrawal in 5 individuals. Seizure outcome was predicted by a combination of the following descriptors: age at epilepsy onset, epilepsy duration, long-term invasive EEG, and specific MRI, and PET findings.

**Significance:** This study highlights the broad phenotypic spectrum of FCD type 1, which spans far beyond the narrow descriptions of previous studies. Combining advanced MRI protocols with additional neuroimaging techniques helped localise the epileptogenic zone in many previously non-lesional cases. Complex multimodal presurgical approaches (including SEEG) could enhance postsurgical outcomes in these complex patients.

**Key points:**

- The phenotypic spectrum of paediatric patients with FCD type 1 spans beyond the narrow description of previous studies
- MRI-negative patients benefit from enhanced precision in localising the epileptogenic zone, facilitated by FDG-PET, SISCOM, and SEEG
- A complex multimodal presurgical approach could enhance postoperative seizure outcomes in patients with FCD type 1
- Paediatric patients with suspected FCD type 1 should be referred to epilepsy surgery centres as soon as possible

## 1. Introduction

Focal cortical dysplasia (FCD) is the most common cause of drug-resistant epilepsy (DRE) in children (1, 2). FCD type 1 is still an enigmatic entity compared with FCD type 2, which has well-described neuroimaging (3), electrophysiological findings (4, 5, 6), and genetic background (7, 8, 9).

Over 40% of patients with MRI-negative epilepsy indicated for resective epilepsy surgery have histopathologically confirmed FCD type 1 (10, 11, 12). FCD type 1 lacks specific radiological hallmarks and does not show any characteristic interictal electroencephalogram (EEG) features on scalp EEG (13). However, a study (14) recently described differences in some characteristics of interictal epileptiform discharges (IEDs) and repetitive discharges (RDs) in intracranial EEG recordings between patients with FCD types 1 and 2.

Neurodevelopmental delay is a significant comorbidity in patients with FCD; approximately 53–68% of patients have cognitive impairment (15, 16). Patients with FCD 1 have significantly lower developmental quotient (DQ)/intelligence quotient (IQ) scores than those with FCD type 2 (17). Other neuropsychological comorbidities, such as autism spectrum disorder (ASD), are more likely to be associated with FCD type 1 than with FCD type 2 (18). Cognitive decline in epilepsy is a common characteristic of FCD type 1 (19). Because of the recurrent seizures and neuropsychological comorbidities, epilepsy has a devastating impact on quality of life in patients with FCD type 1 (20).

Epilepsy surgery represents the only effective treatment for FCD type 1 (21), but most studies have reported that the postsurgical seizure outcomes are significantly worse than those for FCD type 2 (22) or long-term epilepsy-associated tumours (23). Previous studies have reported postsurgical seizure freedom in 21% (24) to 47% (25) of patients with FCD type 1.

This study comprehensively characterised the clinical phenotype of paediatric patients with isolated FCD type 1 to expand the understanding of the phenotypic spectrum of these patients. Additionally, we conducted a systematic re-analysis of electrophysiological and radiological features and outcomes and their predictors.

## 2. Materials & methods

### 2.1. Patient selection

This study involved a cohort of paediatric patients with focal DRE (26) who underwent resective or disconnective epilepsy surgery at Motol University Hospital in Prague between 2010 and January 2023. Included patients fulfilled the following criteria: 1) histopathological diagnosis of isolated FCD type 1 according to the latest International League Against Epilepsy (ILAE) classification (27) and 2) at least 1 year of postsurgical follow-up. Two expert neuropathologists (JZ and MK) re-evaluated all brain specimens to reach a consensus on FCD type 1 and its subtypes.

### 2.2. Clinical history; neurological and neuropsychology assessments

Individual patient characteristics obtained from electronic health records were retrospectively collected and re-analysed with a focus on epilepsy phenotype and neuroimaging and electrophysiological features, which served as independent descriptors for subsequent statistical analyses. Epileptic seizures were classified according to the ILAE classification (28). To characterise the time course of epilepsy, the following intervals were defined: 1) duration from epilepsy onset to epilepsy surgery and 2) duration from the first examination in our centre to surgery.

Patients underwent neuropsychological assessment in the presurgical evaluation and 1 year after surgery. The following age-dependent tests were used: the Bayley Scale of Infant and Toddler Development, 3rd edition (BSID-III) (29); the Wechsler Preschool and Primary Scale of Intelligence, 4th edition (WPPSI-IV) (30); the Wechsler Intelligence Scale for Children, 3rd edition (WISC-III) (31); and the Wechsler Adult Intelligence Scale, 3rd edition (WAIS-III) (32).

### 2.3. Electrophysiology

All patients underwent scalp video EEG monitoring, encompassing wakefulness and sleep states for a minimum duration of 24 hours; at least two habitual epileptic seizures were captured. A systematic re-analysis of the EEG characteristics was conducted by PK with a focus on 1) background activity (normal or slow), 2) sleep architecture (normal or disrupted) and the presence of sleep spindles (symmetric, asymmetric, or absent), 3) local slowing (not present, regional, multiregional, hemispheric, or generalised), 4) the extent and localization of interictal epileptiform discharges (IEDs) (not present, regional, multiregional, hemispheric, or generalised), 5) the presence of secondary bilateral synchrony (bilateral spread of IEDs), 6) the presence of EEG status epilepticus (continuous IEDs), and 7) the extent and localization of ictal activity (not lateralised, regional, multiregional, hemispheric, or generalised).

### 2.4. Neuroimaging

All patients underwent high-resolution brain MRI (1.5T or 3T) using a dedicated epilepsy MRI protocol (33). Two experienced paediatric neuroradiologists (MK and ZH) performed a systematic re-evaluation of all MRI studies, including consensus on the presence or absence of the following five radiological features defined in previous radiological studies on patients with FCD (25, 34, 35, 36): 1) increased signal in the white matter on T2 and FLAIR; 2) hypoplasia of the white matter; 3) blurring of the grey–white-matter boundary (GWMB); 4) cortical thickening; and 5) abnormal gyral pattern. MRI findings were classified as focal, lobar, multilobar, or hemispheric, and the localisation of the MRI abnormalities was specified. The specifications of the MRI protocols are provided in Supplementary Table 1.

All patients underwent fluorodeoxyglucose positron emission tomography (FDG-PET) co-registered with MRI. For this study, images were re-analysed and postprocessed by partial volume effect correction (37) to highlight cortical metabolic abnormalities, which were subjectively labelled as obvious hypometabolism, subtle or moderate hypometabolism, normal metabolism, and hypermetabolism, and compared with resection: concordant, overlapped, partially overlapped, and discordant. In selected cases, ictal and interictal single-photon emission computed tomography (SPECT) (99mTc-ECD and subsequently 99mTc-HMPAO) had been performed and processed into subtraction ictal SPECT co-registered to MRI (SISCOM). The SISCOM findings were classified as focal, lobar, multilobar, or non-localised and compared with resection: concordant and discordant. A description of FDG-PET is provided in Supplementary 1.

All lesions and resection extents were manually outlined by ZH and KM, respectively, in the preoperative MRI using 3D Slicer software and exported to binary label maps with voxels belonging to the lesion/resection (labelled 1) and background (labelled 0). The lesion/resection volumes were computed as the number of voxels (labelled 1) multiplied by the volume of an individual voxel. A detailed description of the volumetric analysis is provided in Supplementary 2.

### 2.5. Intracranial EEG

Long-term intracranial video/EEG monitoring (iEEG) was indicated if 1) the patient had non-localising findings on MRI or discordant findings in non-invasive examinations or 2) the structural abnormality was adjacent to eloquent cortical areas. The implantation scheme of the depth (stereoelectroencephalography, SEEG) electrodes (DIXI medical, Besançon, France) was planned with a neuronavigation system (StealthStation S7, Medtronic, Dublin, Ireland) for frame non-robot Cosman–Roberts–Wells stereotaxy (Integra LifeSciences, Princeton, New Jersey, USA). Early cases (n = 2) underwent implantation with subdural electrodes (Integra LifeSciences). Post-implantation computed tomography (CT) was fused with pre-surgical MRI images to obtain recording contact positions related to brain anatomy (38).

### 2.6. Epilepsy surgery, completeness of resection, and outcomes

The localisation of the epileptogenic zone (EZ), estimated using seizure semiology and electrophysiological (iEEG and stimulation mapping) and neuroimaging (MRI, FDG-PET, and SISCOM) findings, was based on expert panel consensus. The completeness of resection was computed as the proportion of the MRI lesion volume included in the resection using the label maps of the outlined lesions/resections. In MRI-negative/inconclusive cases, the completeness was determined by the proportion of iEEG channels within seizure onset that were included in the resection. The completeness of hemispheric disconnection was evaluated by the analysis of postsurgical MRI and diffusion tensor imaging (DTI) tractography.

Postsurgical seizure outcomes were assessed using the ILAE scale (39) with a 2-year follow-up (except for the last two cases, for which the follow-up was 1 year). Data regarding anti-seizure medications (ASMs) were evaluated as follows: 1) a reduction in the number of ASMs (at least one ASM withdrawn), 2) complete discontinuation of ASMs, and 3) unchanged or modified without a significant reduction.

### 2.7. Statistical analysis and modelling

Twenty-eight descriptors of clinical history, electrophysiological, neuroimaging, and neuropsychological findings, and seizure outcomes were selected to represent the patient cohort. Considering the limited sample size, descriptive statistics were predominately used to characterise the cohort. To complement the descriptive characteristics, stepwise generalised linear models (GLMs) with a binomial distribution and the logit link function were used to identify possible combinations of descriptors associated with favourable (ILAE 1) or unfavourable (ILAE 2–5) seizure outcomes. To reduce the model variance and eliminate the collinearity of model predictors, only a subset of descriptors was selected to build the GLMs. In cases of correlation or dependence (p < 0.05) between a pair of descriptors, only one representative descriptor with higher clinical importance was included in the GLM. The descriptor selection process revealed interesting dependencies between descriptors, some of which are reported in the text. All analyses were performed with MATLAB software (ver. 2023b, MathWorks, US). Further details are provided in Supplementary 3.

## 3. Results

### 3.1. Clinical characterisation

We enrolled 31 paediatric patients (17 females and 14 males); 27 patients (87.1%) had histopathologically confirmed FCD type 1A and 4 patients (12.9%) had FCD type 1C. The clinical data of included patients are summarised in Table 1. None of the children had febrile seizures. Three patients (9.7%) had a family history of epilepsy (1st or 2nd degree). Ten patients (32.3%) exhibited abnormal focal or non-focal neurological findings (such as hypotonia). Six patients (19.4%) had perinatal complications (e.g. prematurity, defined by the World Health Organization as any birth before 37 weeks of gestation or a low Apgar score (below 7) at any time point) without neuroradiological abnormalities suggestive of perinatally acquired brain lesions. Developmental delay before epilepsy onset was noted in five patients (16.1%). In 25 patients (80.6%), developmental arrest, regression, or cognitive decline was observed during the course of epilepsy.

The median age of epilepsy onset was 4.2 years (range 0.3–16.0 years), and 15 patients (48.4%) had epilepsy onset before the age of 3 years. Daily seizures were reported in 27 patients (77.4%). The median age at the first examination at the Motol Epilepsy Centre was 8.0 years (range 1.1–17.8 years). The median duration from epilepsy onset to referral to the epilepsy centre was 2.3 years (range 0.1–16.8 years). Various types of focal seizures were reported in 27 patients (87.1%), and epileptic spasms were reported in 7 individuals (22.6%). Sixteen patients (51.6%) experienced focal to bilateral tonic–clonic seizures (FBTCSs). Additionally, four patients (12.9%) had episodes of status epilepticus. All patients developed drug resistance, and the median number of ASMs was 6 (range 2–11). Fifteen patients (48.4%) experienced a period of temporary seizure freedom (defined in this study as a minimum of 3 months without experiencing seizures).

### 3.2. Electrophysiological characterisation

The main electrophysiological features observed in our cohort are summarised in Table 2. Background EEG activity was normal in 17 patients (54.8%). Disrupted sleep architecture was noted in 16 patients (51.6%), and 9 patients (29.0%) exhibited asymmetry or absence of sleep spindles. Localised EEG slowing was regional in 7 patients (22.6%), multiregional in 16 (51.6%), hemispheric in 4 (12.9%), and generalised in 3 (9.7%), and 1 patient showed no localised slowing. Localised EEG slowing was intermittent in 19 patients (60.0%) and continuous in 12 patients (40.0%). Interictal EEG spikes were predominantly multiregional and were noted in 20 individuals (64.5%), with the maximum over the temporal or frontotemporal regions. Ictal EEG patterns were multiregional in 13 patients (41.9%), regional in 8 (25.8%), generalised in 4 (12.9%), hemispheric in 3 (9.7%), and not lateralised in 3 (9.7%). Secondary bilateral synchrony was noted in 13 patients (41.9%). Only one patient had regional continuous epileptiform discharges on interictal EEG.

### 3.3. Neuropsychological characterisation

Presurgical IQ scores were available for 28 out of 31 patients, and the median score was 76 (range 32–106). The descriptor selection process revealed interesting trends related to neuropsychological characterisation. Lower scores were correlated with an earlier age of epilepsy onset (r = 0.4; p = 0.03, Spearman correlation). Patients with normal sleep architecture (n = 16) had higher presurgical IQ scores than those with disrupted sleep architecture (n = 12) (p = 0.04, Kruskal–Wallis) (84; range 43–106 vs. 68; range 32–99). Similarly, patients (n = 17) with normal background EEG activity had higher IQ scores than patients with slow background EEG activity (n = 11) (p = 0.02, Kruskal–Wallis) (85; range 56–105 vs. 64; range 32–106).

Among other neuropsychological comorbidities, two patients had attention deficit hyperactivity disorder (ADHD) and three patients with ASD, one of whom had Asperger syndrome.

### 3.4. Neuroimaging characterisation

Twenty-three patients (74.2%) had visible MRI abnormalities, whereas eight patients (25.8%) displayed no signs of malformation of cortical development. Initial MRI (taken and evaluated at our centre) was reported as normal or abnormal without signs of FCD (e.g. non-specific gliosis) in 24 patients (77.4%), but MRI performed with a dedicated epilepsy protocol at our centre revealed signs of FCD in 17 of 24 initially negative patients (70.8%). The left hemisphere was affected in 14 patients who were MRI-positive (60.9%). Table 3 summarises the radiological features observed in our cohort.

The most common radiological features observed in the MRI-positive subgroup were an increased white matter signal in T2W and FLAIR sequences (95.7%), hypoplasia of the white matter (87.0%), and blurred grey–white matter junction (82.6%). Cortical thickening (47.8%) and abnormal gyral patterns (47.8%) were less common. The extent of MRI findings was lobar in 11 patients (47.8%), multilobar in 11 cases (47.8%), and hemispheric in 1 case (4.3%).

SISCOM was conducted in 18 patients (58.1%) with focal (n = 1), lobar (n = 5), multilobar (n = 6), and non-localised (n = 6) findings. The SISCOM finding was concordant with the resection in 10 of the 18 patients (56%) and discordant in 8 of the 18 patients (44%). All 31 patients in the study underwent FDG-PET. An obvious hypometabolism on FDG-PET was observed in 17 patients (54.8%), whereas a subtle or moderate hypometabolism was observed in 8 patients (25.8%). Three patients (9.7%) had normal metabolism on FDG-PET. Localised hypermetabolism was observed in three individuals (9.7%). The identified abnormal region was concordant with the resection in 8 patients (25.8%), overlapped with the resection in 4 patients (12.9%), and partially overlapped in 14 patients (45.2%). In the MRI-negative subgroup (n = 8), a combination of FDG-PET and SISCOM helped localise the epileptogenic zone in 6 of the 8 patients (75.0%). Only one patient had non-localised FDG-PET and SISCOM.

### 3.5. Epilepsy surgery and completeness of resection

The median age at epilepsy surgery was 9.3 years (range 1.5–20.0 years), and the median duration of epilepsy was 3.4 years (range 1.0–17.8 years). The median duration from the first examination at our centre to epilepsy surgery was 11 months (range 3–51 months). Sixteen patients (51.6%) underwent long-term iEEG monitoring before epilepsy surgery; half of them were MRI-negative. Twenty-eight patients (90.3%) underwent resective surgery. The extent of resection was focal (sublobar) in 8 patients (25.8%), lobar in 10 patients (32.3%), and multilobar in 10 patients (32.3%). Three patients (9.7%) underwent hemispheric disconnection. Reoperation was necessary in one patient.

The median volume of resection (excluding hemispherectomy) was 38 cm^3^ (range 8–129 cm^3^), and the resection extent was similar in MRI-lesional and MRI-negative groups. The median completeness of lesion resection as assessed with MRI was 77.0% (range 48.0–100%); the completeness of resection as assessed with iEEG was 88.2% (range 21.4–100%). Complete hemispheric disconnection was achieved in all three patients.

The multimodal approach resulting in resective surgery in selected cases is illustrated in Figure 1.

**Figure 1:**
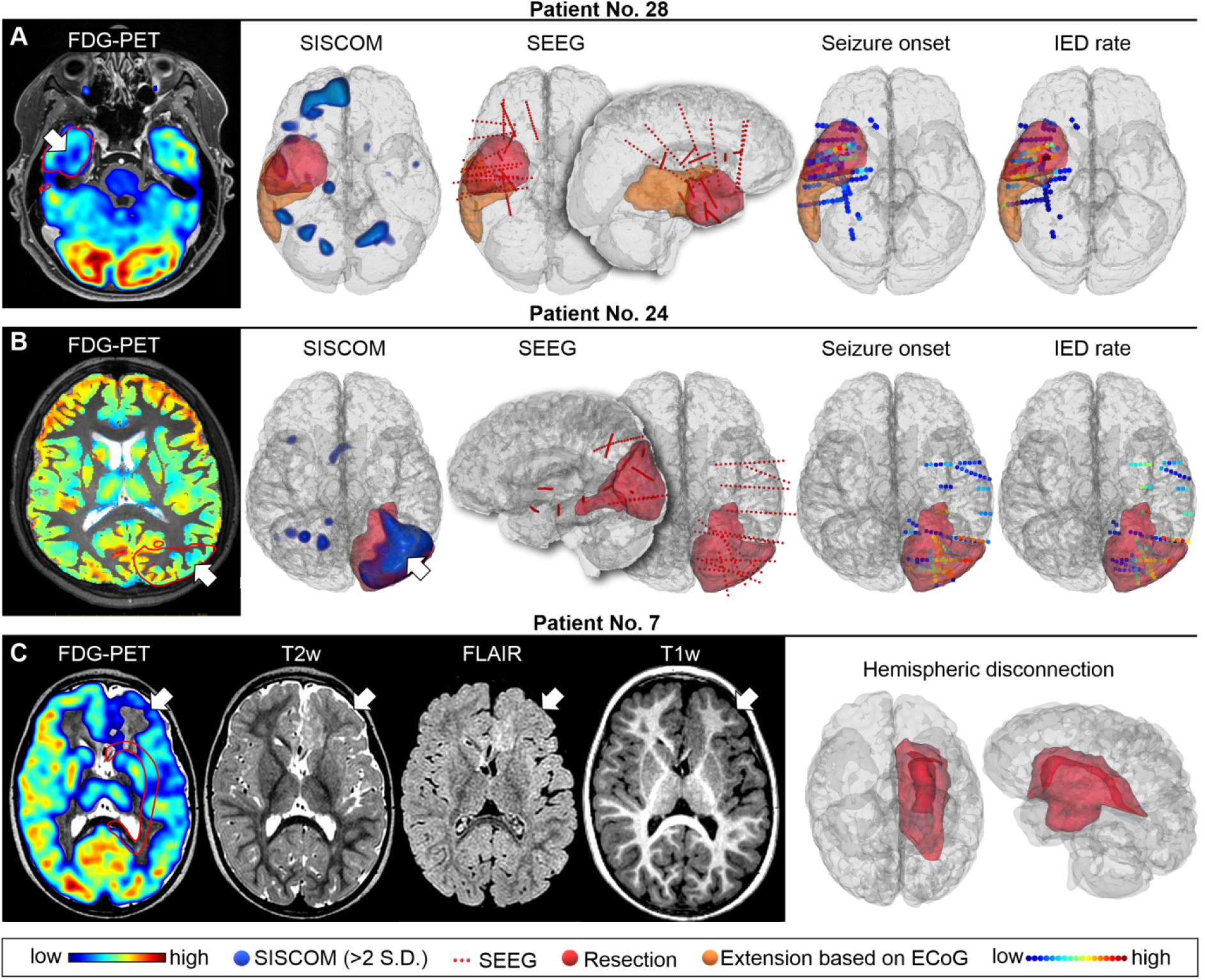
An example of the importance of multimodal assessment in three postoperative seizure-free patients. (A) Patient 28 had normal magnetic resonance imaging (MRI) findings and lateralising but non-localising single-photon emission computed tomography co-registered to MRI (SISCOM). Scalp EEG showed extensive frontotemporal epileptiform activity on the right. Fluorodeoxyglucose positron emission tomography (FDG-PET) with additional postprocessing revealed a mild hypometabolism on the base of the temporal pole (arrow), which was the target of stereoelectroencephalography (SEEG) exploration. Seizure onset zones (48) were confirmed in the hypometabolic tissue. However, interictal epileptiform discharges (IEDs) were widespread in the temporal lobe, indicating an extensive epileptic network (49). The initial resection plan (red area) was extended intraoperatively on the basis of electrocorticography (ECoG) (orange area) including the region uncovered by SEEG electrodes. The patient was completely seizure-free during follow-up. (B) Patient 24 had normal MRI findings with subtle hypometabolic abnormalities in FDG-PET on left parieto-occipital lobes (arrow) and multilobar left frontotemporal EEG findings. SISCOM localised the epileptogenic zone to the left temporo-parieto-occipital region (arrow), which was confirmed by SEEG. (C) Patient 7 (an infant) had extensive left-sided structural abnormalities visible on MRI in the frontoinsular-temporal region with the corresponding FDG-PET hypometabolism (arrows). The patient underwent a left hemispherectomy.

### 3.6. Outcomes and analysis of their predictors

By the last follow-up, 22 patients (71.0%) had achieved seizure freedom (ILAE class 1). Two patients (6.5%) were classified as ILAE class 3, 4 patients (12.9%) were classified as ILAE class 4, and 3 patients (9.7%) were classified as ILAE class 5. Notably, in the MRI-negative subgroup, 7 of the 8 patients (87.5%) achieved long-term seizure freedom. The seizure outcome was similar in the MRI-negative and MRI-positive groups (p = 0.23, Chi-squared test). ASMs were reduced in 21 patients (67.7%), and complete withdrawal of ASMs was possible in 5 individuals (16.1%). Patients with regional ictal EEG patterns had a better chance of reducing ASMs (p = 0.04, Chi-square test).

Neuropsychological profiles before surgery and after 1 year of follow-up were available in 24 patients (77.4%). The median postsurgical IQ score was 71 (range 21–110). We did not observe a correlation between a postsurgical increase or decrease in IQ and seizure outcomes. Three patients exhibited an increase in IQ score of over 10 points (one was seizure-free, one was in ILAE class 3, and one was in class 5), and three exhibited a decrease in IQ score of over 15 points (two were seizure-free; one was in ILAE class 5).

Seizure outcome class was predicted by a GLM with a combination of the following descriptors: age at epilepsy onset (A), epilepsy duration (D), performance of iEEG exploration (I), extent of MRI (M), and PET (P) findings. The resulting GLM model was as follows: logit(ILAE class) = 1.63 − 0.05A − 0.03D + 2.6I + 3.28P − 2.34M; p < 0.003. The 95% confidence intervals for the coefficient estimates were (−4.557, 7.811) for the intercept, (−0.108, 0.002) for A, (−0.065, 0.005) for D, (−0.748, 5.938) for I, (0.088, 6.462) for P, and (−5.497,0.824) for M. This model suggests that earlier epilepsy onset is associated with a higher probability of a favourable outcome, and iEEG monitoring increased this probability in patients with late epilepsy onset. Shorter epilepsy duration, less extensive MRI, and clearly visible PET abnormalities increased the probability of seizure freedom. The detailed results are provided in Supplementary 4.

### 3.7. Long-term experience at Motol Epilepsy Centre

The descriptor selection process revealed interesting trends related to long-term experience at Motol Epilepsy Centre. Although the resection volume decreased with time (r = −0.54; p < 0.003, Spearman correlation), the resection completeness improved (r = 0.41; p = 0.03, Spearman correlation). The duration of epilepsy decreased with time (r = −0.41; p = 0.02, Spearman correlation), whereas the duration from the first examination at our centre to epilepsy surgery remained the same. The effect of the year of surgery on the seizure outcome was not observed. Twenty patients (64.0%), including 7 of the 8 patients in the MRI-negative subgroup (88.0%), were indicated for epilepsy surgery after 2017.

## 4. Discussion

FCD type 1 is one of the most challenging causes of focal DRE in surgical cohorts; it is always associated with terms “difficult to diagnose” or “difficult to treat”. To the best of our knowledge, this study presents the largest single-centre cohort of paediatric patients with histopathologically confirmed FCD type 1, comprising 31 children. We focused on re-analysing electro-radiological characteristics, describing clinical features in detail, and analysing outcome predictors.

Our data suggest extreme phenotypic variability, expanding the classical phenotype of FCD type 1 patients described by Holthausen and colleagues (13, 24) with the characteristic of age at epilepsy onset in infancy or early childhood. Despite the predominance of FCD type 1A in our cohort, the clinical phenotype does not completely overlap with the recently described disease multilobar unilateral hypoplasia with severe epilepsy in children (MUHSEC), which is characterized by early-onset drug-resistant epilepsy, severe mental retardation, and characteristic electrophysiological and neuropathological findings (40). The age of epilepsy onset in our cohort was highly variable, and these results are concordant with the findings reported by Yao 2016 (41) and Guerrini 2021 (42). None of our patients exhibited neonatal seizures or epilepsy onset in the first 2 months of life; this is comparable to MUHSEC and differs from extensive FCD type 2 (40). Most patients in our study experienced daily seizures, similar to patients with MUHSEC; however, some patients experienced weekly or monthly seizures, similar to patients in a recent study (22). The median duration of epilepsy in our study was 3.4 years, which is slightly below the duration reported in the large European multicentre epilepsy surgery series (43). We observed a shorter duration of epilepsy in patients who underwent surgery in later years. We believe that this is a result of nationwide education, which shortened the time until referral to an epilepsy centre.

We observed a negative impact of ongoing epilepsy on children’s development and cognition in most of our cases, consistent with previous studies (15, 16). Limited evidence indicates that focal epilepsy disrupts the structure and function of sleep spindles, which may play a role in impaired cognitive functioning (44). However, we did not observe a difference in presurgical IQ scores between patients with asymmetric or absent and symmetric sleep spindles. Interestingly, we observed that slow background EEG activity and disrupted sleep architecture were linked to lower presurgical IQ scores. This suggests that extensive epileptiform activity may partly contribute to abnormalities in sleep microarchitecture, affecting cognitive and behavioural functioning beyond the direct impact of seizures (45). We did not observe a significant difference between presurgical and postsurgical IQ scores, possibly because of the short follow-up period.

Interictal and ictal EEG findings frequently suggested multiregional epileptiform activity with predominance over the temporal, frontal, and parietal regions and a tendency towards generalisation, as evidenced by the phenomenon of secondary bilateral synchrony. Inconsistent with MUHSEC, no patients showed posterior predominance. Extensive epileptiform activity was observed not only in patients with multilobar MRI abnormalities but also in those with lobar distribution and normal MRI findings. These observations suggest that scalp EEG has limited localisation value; thus, more advanced techniques, especially neuroimaging, are required. In contrast to FCD type 2, which is characterised by focal rhythmic epileptiform discharges (4), we did not detect any distinctive EEG features in FCD type 1.

Approximately 60% of patients with FCD type 1 have visible MRI abnormalities (46). However, the utilisation of MRI protocols for epilepsy enabled the identification of FCD-related abnormalities in many of our patients (74.2% of cases). Moreover, the advanced MRI protocols provided at our epilepsy centre revealed FCD features in 70.8% of patients who were classified as MRI-negative when referred to our centre. We observed increased white matter signal intensity in the T2W and FLAIR sequences and hypoplasia of the white matter, which are MRI hallmarks of MUHSEC; additionally, a blurred grey–white matter junction was a frequent feature in our cohort, but this is rare in MUHSEC (40). The distribution of MRI abnormalities was mainly multilobar or lobar, with predominance in the temporal, frontal, and insular lobes. In MRI-negative patients and cases with visible lesions not corresponding to the distribution of epileptiform activity on EEG, additional neuroimaging techniques, such as FDG-PET and SISCOM, can be highly beneficial. Using these examinations, we were able to localise the presumed epileptogenic zone in six of the eight MRI-negative patients. Advances in neuroimaging and the growing experience of our team resulted in an increased number of candidates with FCD type 1 identified for surgical treatment. Two-thirds of patients in our cohort (including most MRI-negative patients), underwent surgery in the second half of the study period (2017–2023).

Surgical resection or disconnection procedures successfully eliminated seizures (ILAE class 1) in 71% of the children. These outcomes are more favourable than those reported in previous studies involving patients with FCD type 1 (22, 24, 25, 40, 47) and are more similar to the postsurgical outcomes of patients with FCD type 2 (22). Notably, MRI-negative patients had excellent seizure outcomes and undoubtedly benefited from enhanced precision in localising the epileptogenic zone facilitated by FDG-PET, SISCOM, and SEEG. Moreover, we observed trends of decreasing resection volume and improving resection completeness in later indicated patients. Analysis of outcome predictors suggests that earlier epilepsy onset, shorter epilepsy duration, less extensive MRI, clearly visible PET abnormalities, and performed iEEG exploration increased the chance of seizure freedom.

## Conclusion

Previous studies (22, 24, 25, 40, 47) have reported unfavourable seizure outcomes in patients with FCD type 1. Our study provides evidence to the contrary—patients with FCD type 1 benefited from epilepsy surgery when it was performed after a complex multimodal presurgical diagnostic process capable of delineating the precise extent of the epileptogenic zone. The phenotypic spectrum of patients with FCD type 1 spans beyond the narrow description of MUHSEC (40) and classical FCD type 1 described by Holthausen and colleagues (13, 24) with varying cognitive and developmental levels, albeit with the universal presence of drug-resistant epilepsy. We recommend that patients with drug-resistant epilepsy with suspected FCD type 1 be referred to epilepsy surgery centres as early as possible.

## Data Availability

Data produced in the present work are contained in the manuscript instead of datails leadet to idnetification of individuals. All data can be available upon reasonable request to the authors.

## Acknowledgements

We confirm that we have read the Journal’s position on issues involved in ethical publication and affirm that this report is consistent with those guidelines.

## Supplements

Supplementary 1 - Volumetric analysis

Supplementary 2 - PET analysis

Supplementary 3 - Statistical analysis

Supplementary Table 1 – Specification of MRI protocols

Supplementary Table 2 – Statistical hypothesis

## Glossary

FCD Type 1 / 2: Focal Cortical Dysplasia Type 1 / 2
DRE: Drug Resistant Epilepsy
ASD: Autism Spectrum Disorder ASM – Antiseizure Medication
IEDs: Interictal Epileptiform Discharges
MRI: Magnetic Resonance Imaging
FDG-PET: Fluorodeoxyglucose-positron Emission Tomography
SPECT: Single Photon Emission Computed Tomography
VEEG: Video Electroencephalography
EEG: Electroencephalography
SEEG: Stereo Electroencephalography
MOGHE: mild MCD with Oligodendroglial Hyperplasia and epilepsy
ILAE: International League Against Epilepsy
SISCOM: Subtraction ictal SPECT coregistered to MRI
FBTCS: Focal To Bilateral Tonic-clonic Seizures
MUHSEC: Multilobar Unilateral Hypoplasia with Severe Epilepsy in Children

## Notes

**Disclosures** None of the authors has any conflict of interest to disclose.

**Funding statement** This work was supported by the Ministry of Health of the Czech Republic - grant projects AZV NU23-04-00209, and AZV NU21-08-00228; by Charles University - grant projects GA UK No 94121 and GA UK No 666320, and by project nr. LX22NPO5107 (MEYS): Financed by EU – Next Generation EU. Katerina Mackova was supported by the Grant Agency of the Czech Technical University in Prague (SGS23/170/OHK3/3T/13). Radek Janca was supported by ERDF-Project Brain Dynamics, No. CZ.02.01.01/00/22_008/0004643. The authors thank CESNET for access to their data storage facility. Alice Maulisova and Katerina Bukacova were supported by the Ministry of Health, Czech Republic - conceptual development of research organisation, Motol University Hospital, Prague, Czech Republic 00064203

### Competing Interest Statement

The authors have declared no competing interest.

### Funding Statement

This work was supported by the Ministry of Health of the Czech Republic - grant projects AZV NU23-04-00209, and AZV NU21-08-00228; by Charles University - grant projects GA UK No 94121 and GA UK No 666320, and by project nr. LX22NPO5107 (MEYS): Financed by EU - Next Generation EU. Katerina Mackova was supported by the Grant Agency of the Czech Technical University in Prague (SGS23/170/OHK3/3T/13). Radek Janca was supported by ERDF-Project Brain Dynamics, No. CZ.02.01.01/00/22_008/0004643. The authors thank CESNET for access to their data storage facility. Alice Maulisova and Katerina Bukacova were supported by the Ministry of Health, Czech Republic - conceptual development of research organisation, Motol University Hospital, Prague, Czech Republic 00064203

### Author Declarations

The study was approved by the institutional Ethical committee of Motol University Hospital, Prague, Czech Republic (2022/06/15 - EK - 602.24/22)

