## Supplementry Table 1, Table 2, and Data for "Multi-layered Diagnostic Protocol Improves Postsurgical Outcomes in Children with Drug-resistant Epilepsy And Focal Cortical Dysplasia Type 1": Supplementary Data.pdf

**Supplementary 1**

Volumetric analysis

MRI lesions were manually outlined by an expert neuroradiologist using multimodal preoperative MRI. If available, planned resection extents were exported from Medtronic StealthStation S7 neuronavigation system and were subsequently compared to postoperative MRI. Potential resection extensions were manually outlined. In case of unavailable neuronavigation data, the resection extends were labeled manually based on preoperative and postoperative MRI.

All MRI lesions and resection extents were outlined in 3D Slicer software and exported to binary label maps with voxels belonging to lesion/resection labeled as 1 and background labeled as 0. The resection volumes were computed using the resection label maps as the number of voxels labeled as 1 multiplied by the volume of an individual voxel.

**Supplementary 2**

PET analysis

All patients had undergone FDG-PET co-registered with MRI. Images were additionally post-processed by partial volume effect correction (38) in MATLAB software (ver. 2023b, Mathworsk, US) with SPM12 image processing toolbox.

FDG-PET images co-registered with MRI together with partial volume effect corrected FDG-PET were displayed in 3D Slicer software. FDG-PET abnormalities were subjectively classified as obvious hypometabolism (clear and well bordered hypometabolism), subtle or moderate hypometabolism (less extensive or moderately decreased hypometabolism), normal metabolism, and hypermetabolism (clear and well bordered hypermetabolism).

The resection binary label map was subsequently added to the 3D Slicer scene. The resection extent was visually compared with the extent of identified FDG-PET abnormalities and classified as concordant (FDG-PET abnormalities agreed with the resection), overlapped (FDG-PET abnormalities were overlapped but more extensive than the resection), partially overlapped (FDG-PET abnormalities were only in part of the resection), and discordant (normal FDG-PET or FDG-PET abnormalities outside the resection).

**Supplementary 3**

Statistical analysis

To complement descriptive characteristics, twenty-eight selected descriptors were investigated to find possible relationships (e.g., between electrophysiological, neuroimaging, and neuropsychological findings), to seizure outcome, phenotype characterization, and long term epilepsy centre experience.

Following descriptors were involved in the statistical analysis: Age at epilepsy onset, preoperative IQ, duration of epilepsy, duration from first examination in our center to surgery, year of surgery, developmental delay before epilepsy onset, developmental regression or cognitive decline during epilepsy, seizure frequency, MRI finding, MRI finding extent, FDG-PET abnormality, FDG-PET abnormality and resection concordance, SISCOM finding, concordance of SISCOM finding and resection, performed iEEG monitoring, background EEG activity, interictal EEG spikes pattern, ictal EEG pattern, concordant ictal and interictal EEG patterns, EEG slowing extent, EEG slowing characteristics, sleep spindles, sleep architecture, resection completeness, resection volume, postoperative IQ, reduced anti-seizure medication, and ILAE outcome class.

Spearman correlation was applied to identify correlated continuous descriptors, the Kruskal-Wallis test to find dependence between continuous and categorical descriptors, and the Chi-square test for categorical descriptors. Due to the limited dataset size, the statistical tests had low power and we expect that multiple-testing correction would yield non-significant results due to type II error. Therefore, we report uncorrected p-values and interpret the results only as trends observed in our cohort that complement descriptive statistics.

A summary of the hypotheses for which the tests yielded significant results (p-value lower than 0.05) is provided in Supplementary Table 2. Only those significant results that were supported by another publication or by clinical observations were reported in the manuscript. The rest of the significant results were likely due to type I error caused by randomness and dataset heterogeneity.

General linear model of seizure outcome

We used stepwise generalised linear models (GLM) with binomial distribution type and logit link function to identify possible combinations of predictors associated with favorable (ILAE 1) or unfavorable (ILAE 2-5) seizure outcome. Only a subset of descriptors was selected to build the GLM to reduce the model variance and eliminate collinearity of model predictors. In case of correlation or dependance ($p<0.05$) between a group of descriptors, only one representative descriptor with the highest clinical importance per group was included in building the GLM. SISCOM descriptors were not included due to the low number of examinations (18/31 patients).

The GLM was built using 9 representative descriptors: age at epilepsy onset, resection completeness, resection volume, duration of epilepsy, ictal EEG pattern, performed iEEG monitoring, FDG-PET abnormalities, FDG-PET abnormalities and resection concordance, and MRI findings extent.

The stepwise predictor selection procedure finally identified 5 predictors contributing to the resulting GLM of seizure outcome: age at epilepsy onset (A), duration of epilepsy (D), performed iEEG monitoring (I), FDG-PET abnormalities (P), and MRI findings extent (M). The resulting GLM was: logit(ILAE class) = $1.63-0.05A-0.03D+2.6I+3.28P-2.34M$; with $p<0.003$. The 95% confidence intervals for coefficient estimates were (-4.557, 7.811) for the intercept, (−0.108, 0.002) for A, (−0.065, 0.005) for D, (−0.748, 5.938) for I, (0.088, 6.462) for P, and (−5.497,0.824) for M. The confidence intervals for model coefficients were considerably wide, most likely due to the limited dataset size. Therefore, we interpret the resulting GLM as a potential model of seizure outcome and suggest that the GLM should be verified on a larger cohort in the future.

#### Supplementary 3, Figure 1

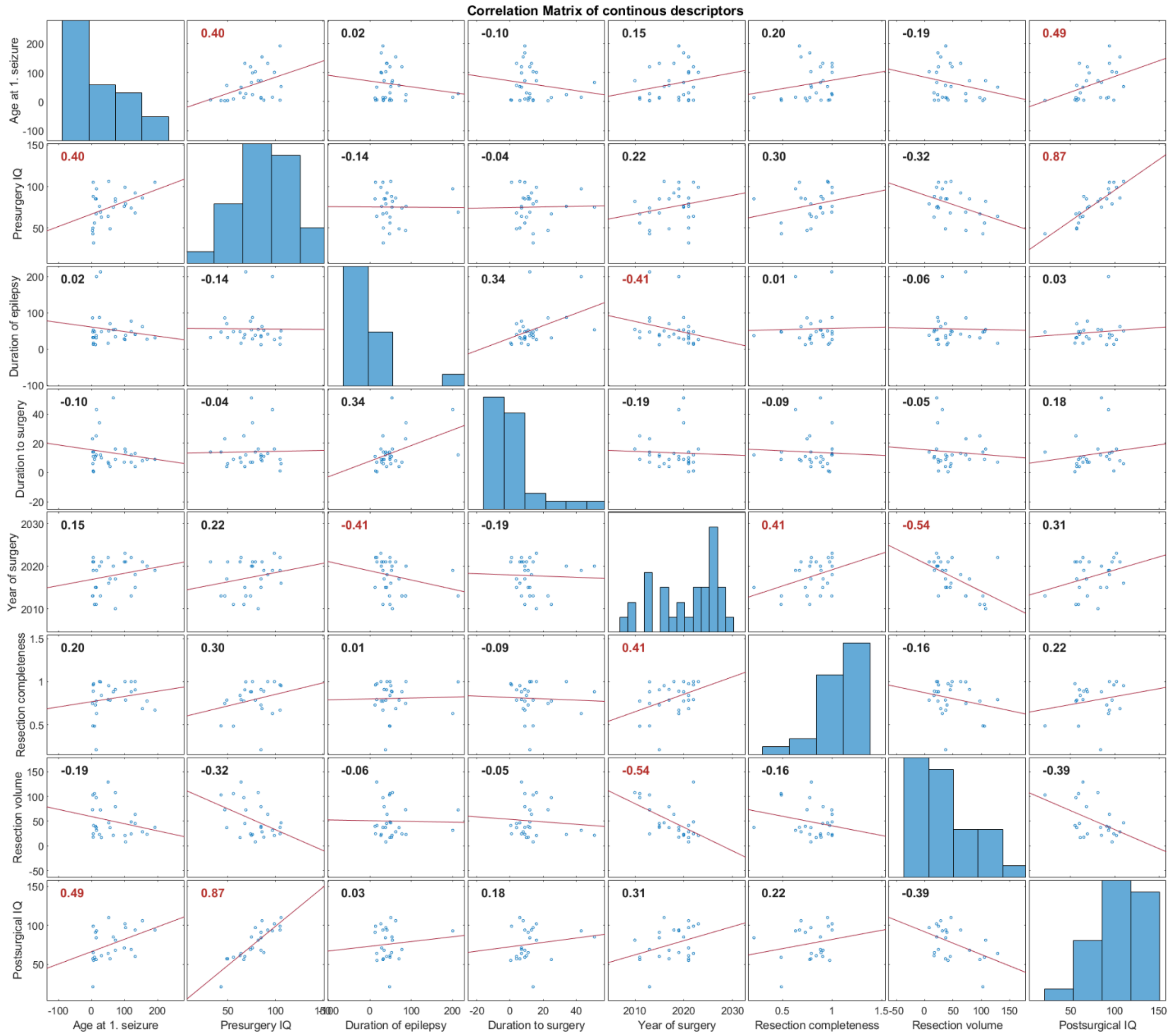


Supplementary Figure 1 shows a correlation matrix of continuous descriptors. Histograms of individual descriptors are displayed on the diagonal. Each subplot shows a correlation between a pair of descriptors; the data points are displayed as blue dots, the red line represents linear fit to the data, and the correlation coefficient is reported in the left upper corner.

#### Supplementary 3, Figure 2

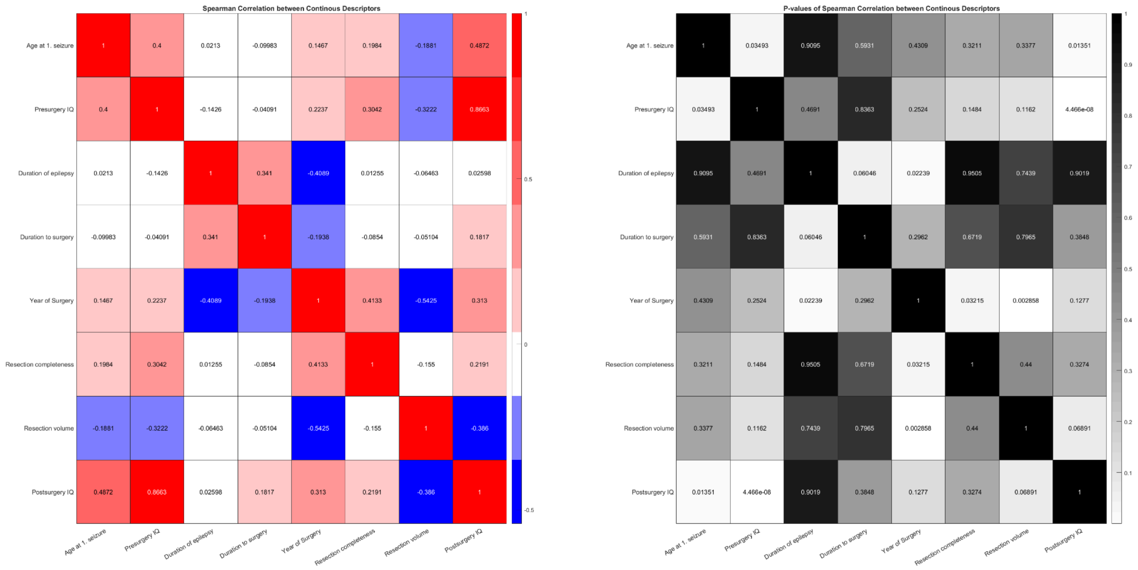


Supplementary Figure 2 shows a matrix of correlation coefficients for all pairs of continuous descriptors (on the left) together with a matrix of correlation p-values (on the right). Positive correlation is displayed in red and negative in blue. Low p-values are displayed in white and high in black.

#### Supplementary 3, Figure 3

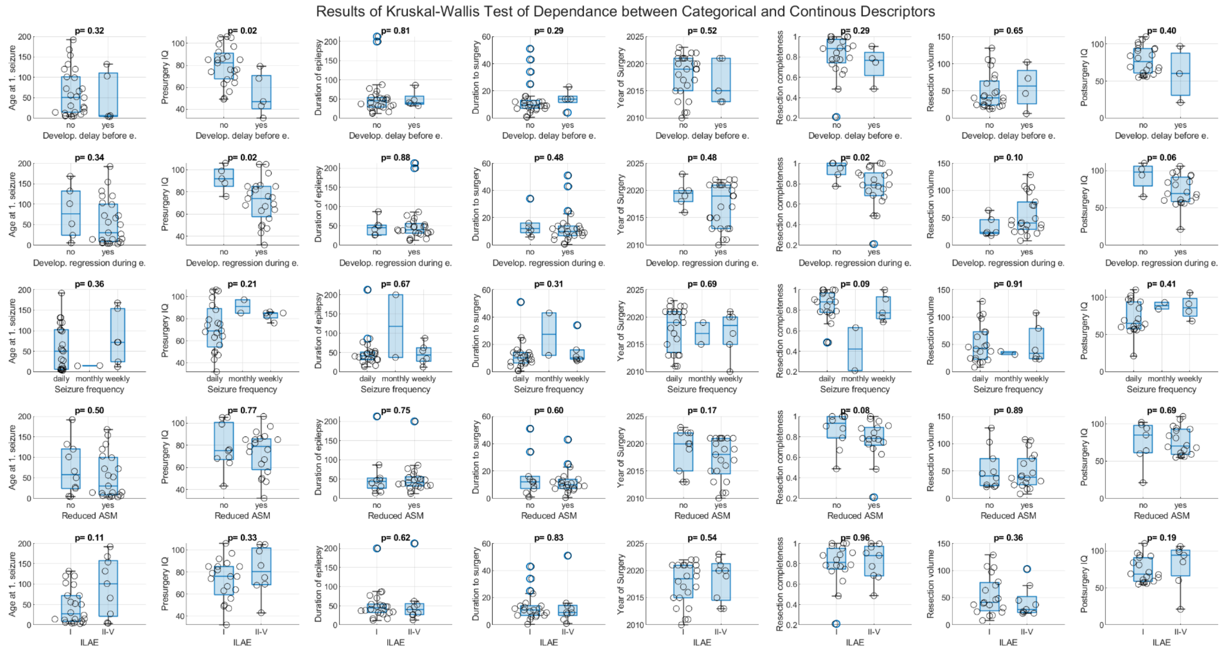


Supplementary Figure 3 shows results of Kruskal-Wallis tests between pairs of categorical and continuous descriptors. In each subplot, the data points are visualised as black circles together with boxplots (blue) and the test p-value is reported in the title.

#### Supplementary 3, Figure 4

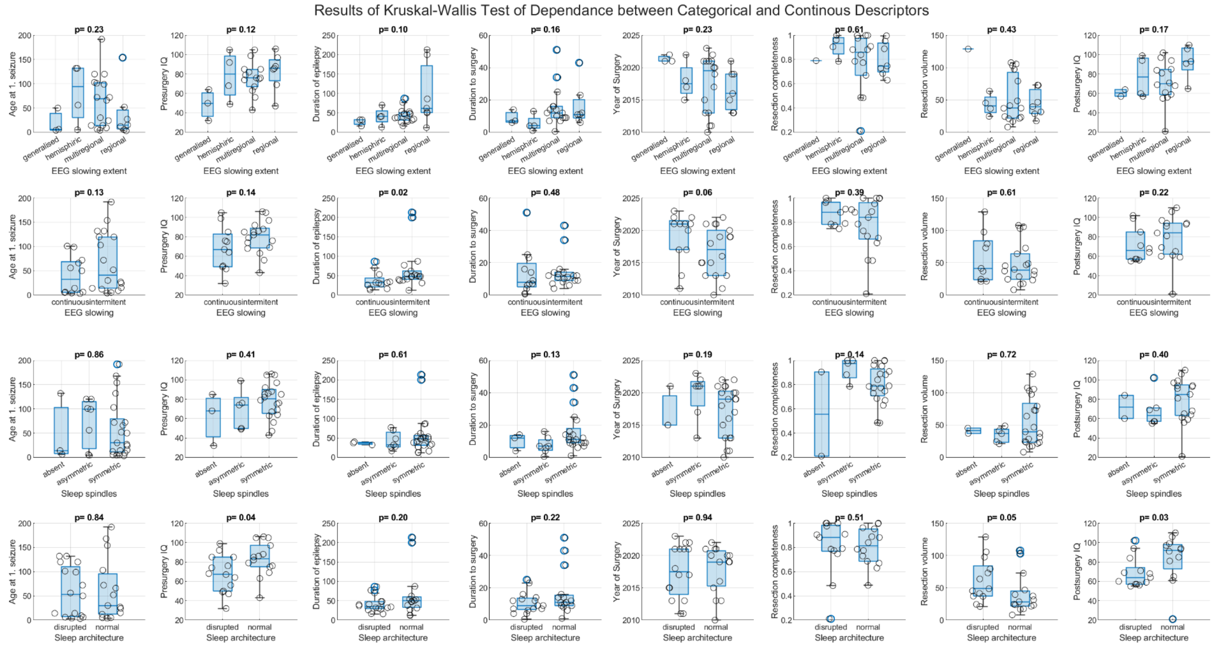


Supplementary Figure 4 shows results of Kruskal-Wallis tests between pairs of categorical and continuous descriptors. In each subplot, the data points are visualised as black circles together with boxplots (blue) and the test p-value is reported in the title.

#### Supplementary 3, Figure 5

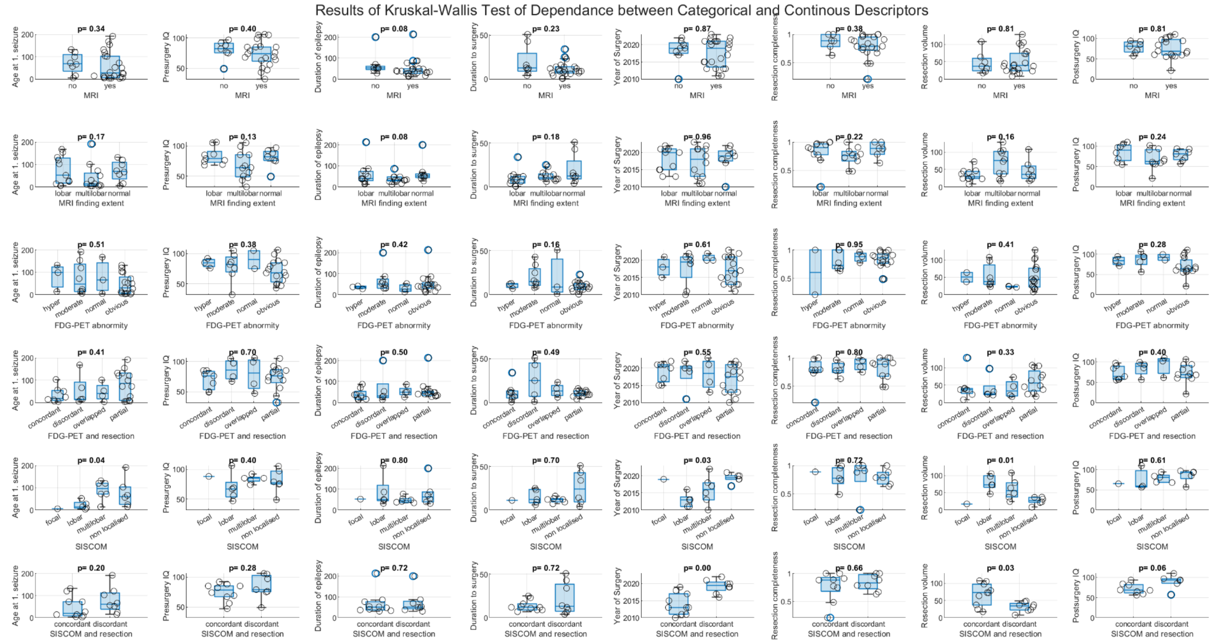

Supplementary Figure 5 shows results of Kruskal-Wallis tests between pairs of categorical and continuous descriptors. In each subplot, the data points are visualised as black circles together with boxplots (blue) and the test p-value is reported in the title.

#### Supplementary 3, Figure 6

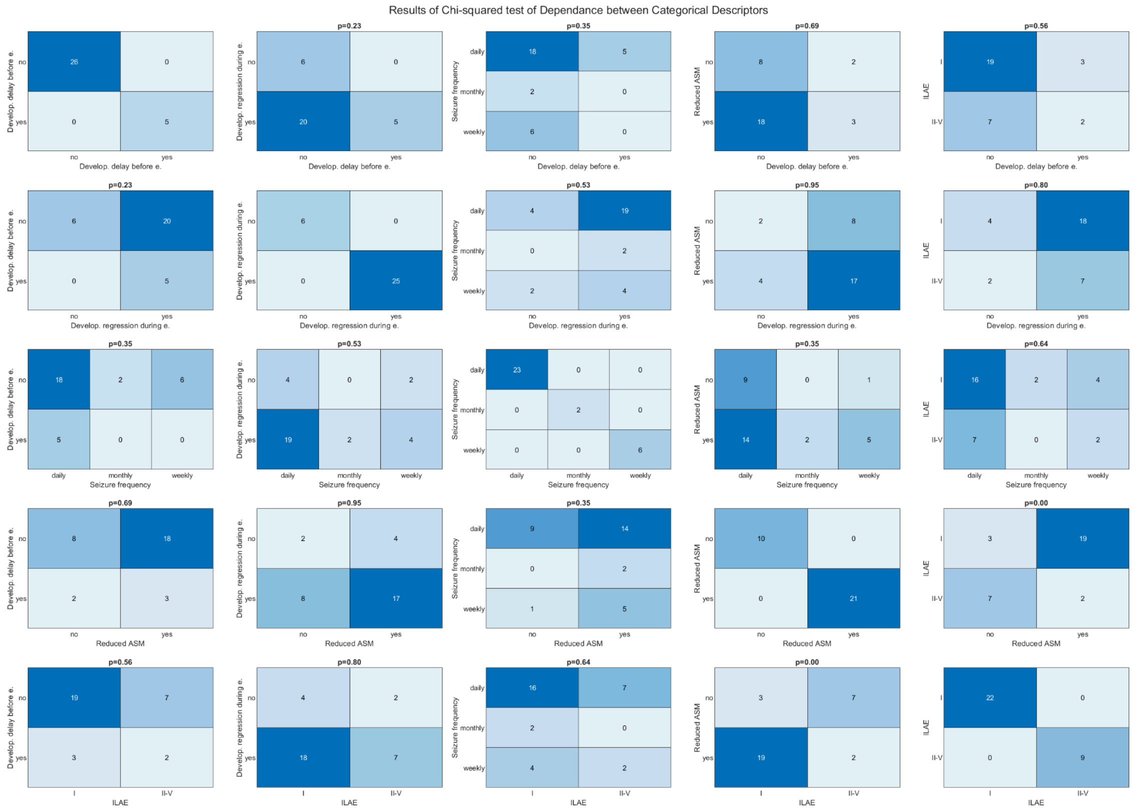


Supplementary Figure 6 shows results of Chi-squared tests between pairs of categorical descriptors. In each subplot, the data are represented by a contingency table and the test p-value is reported in the title.

#### Supplementary 3, Figure 7

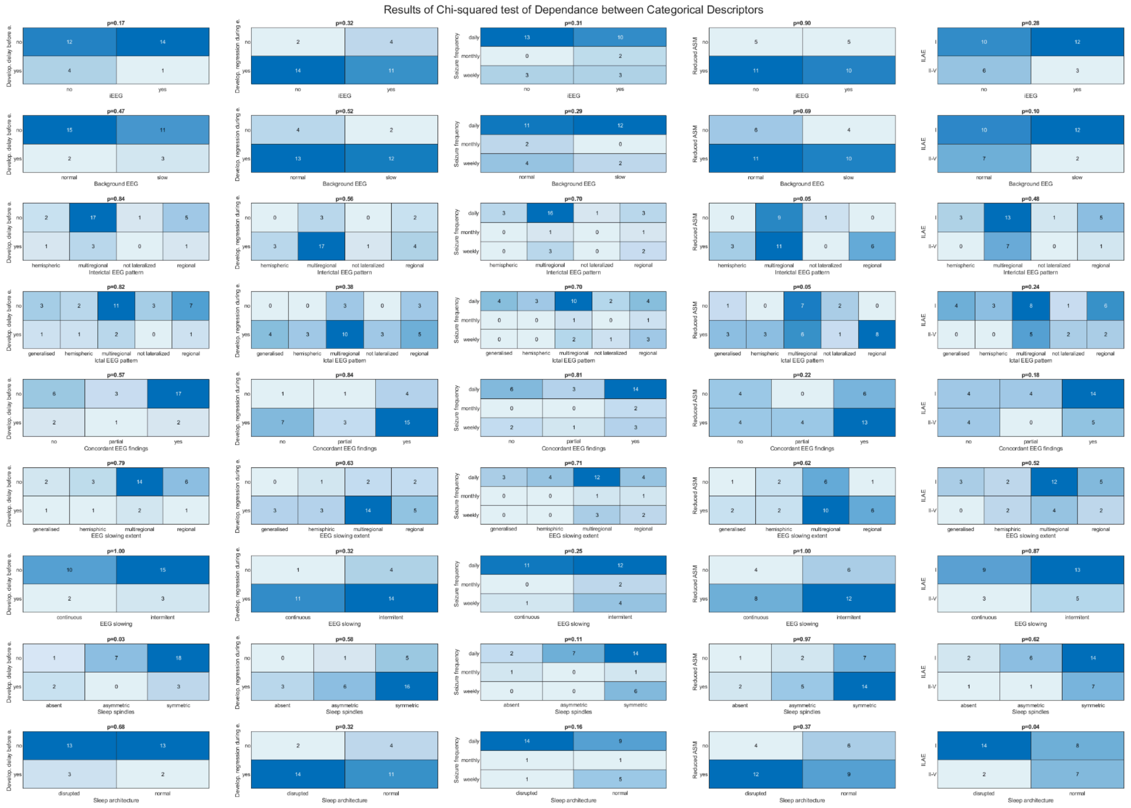


Supplementary Figure 7 shows results of Chi-squared tests between pairs of categorical descriptors. In each subplot, the data are represented by a contingency table and the test p-value is reported in the title.

#### Supplementary 3, Figure 8

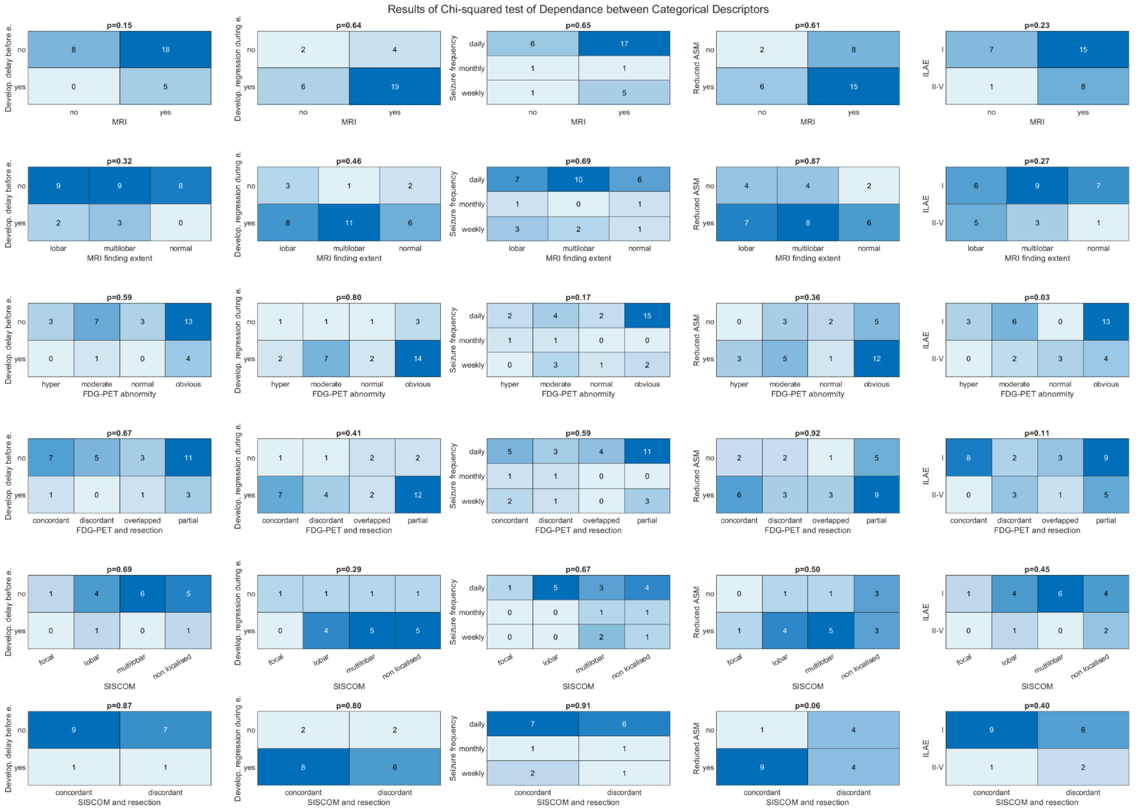


Supplementary Figure 8 shows results of Chi-squared tests between pairs of categorical descriptors. In each subplot, the data are represented by a contingency table and the test p-value is reported in the title.

### Supplementary 3, Figure 9

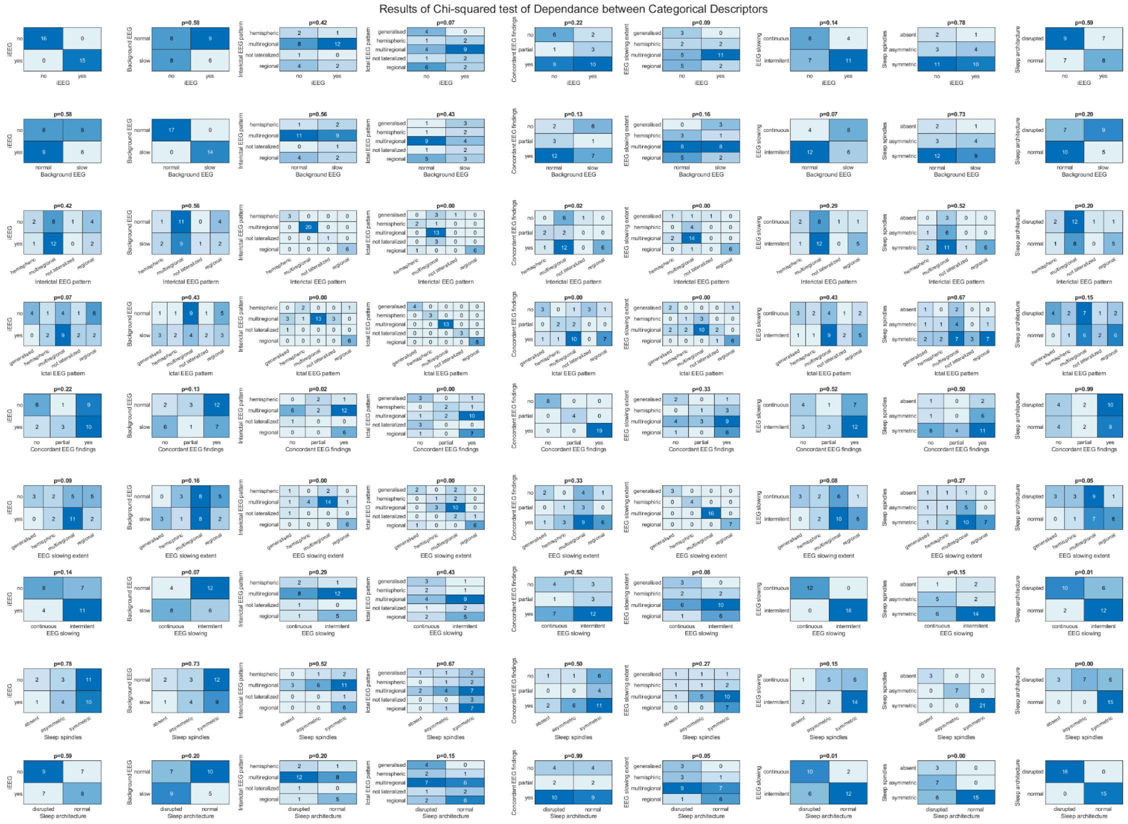

Supplementary Figure 9 shows results of Chi-squared tests between pairs of categorical descriptors. In each subplot, the data are represented by a contingency table and the test p-value is reported in the title.

#### Supplementary 3, Figure 10

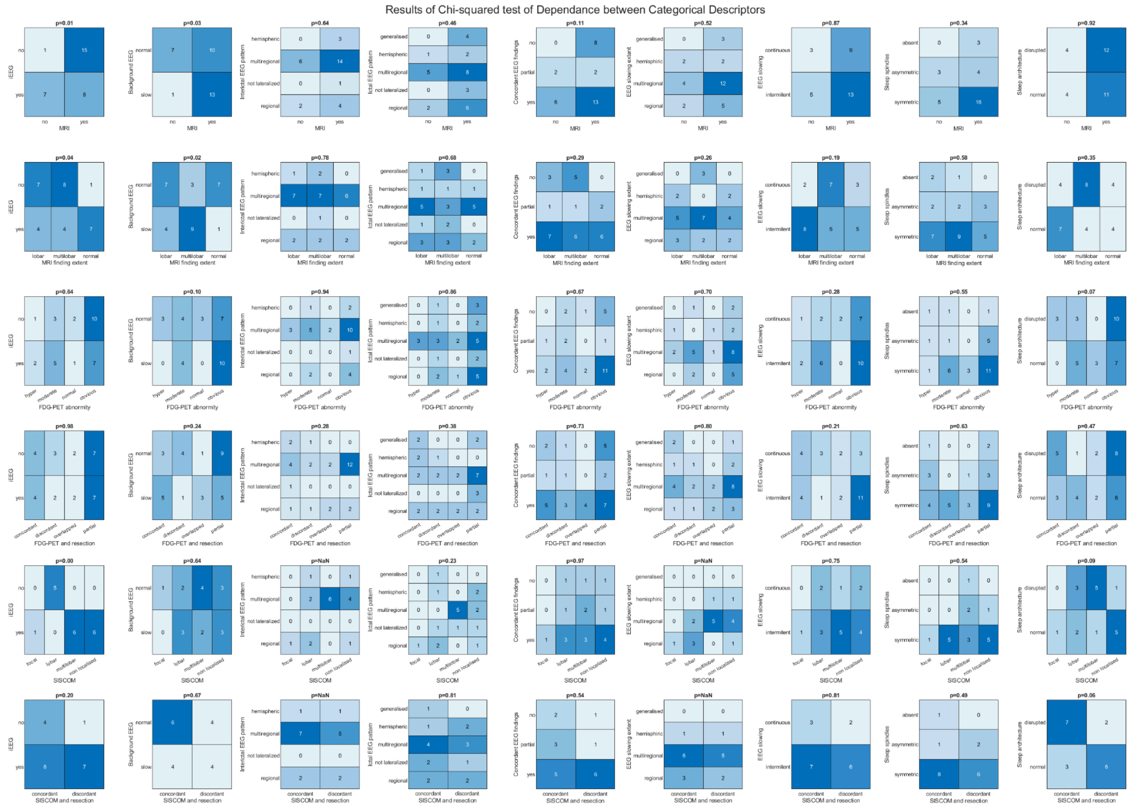


Supplementary Figure 10 shows results of Chi-squared tests between pairs of categorical descriptors. In each subplot, the data are represented by a contingency table and the test p-value is reported in the title.

#### Supplementary 3, Figure 11

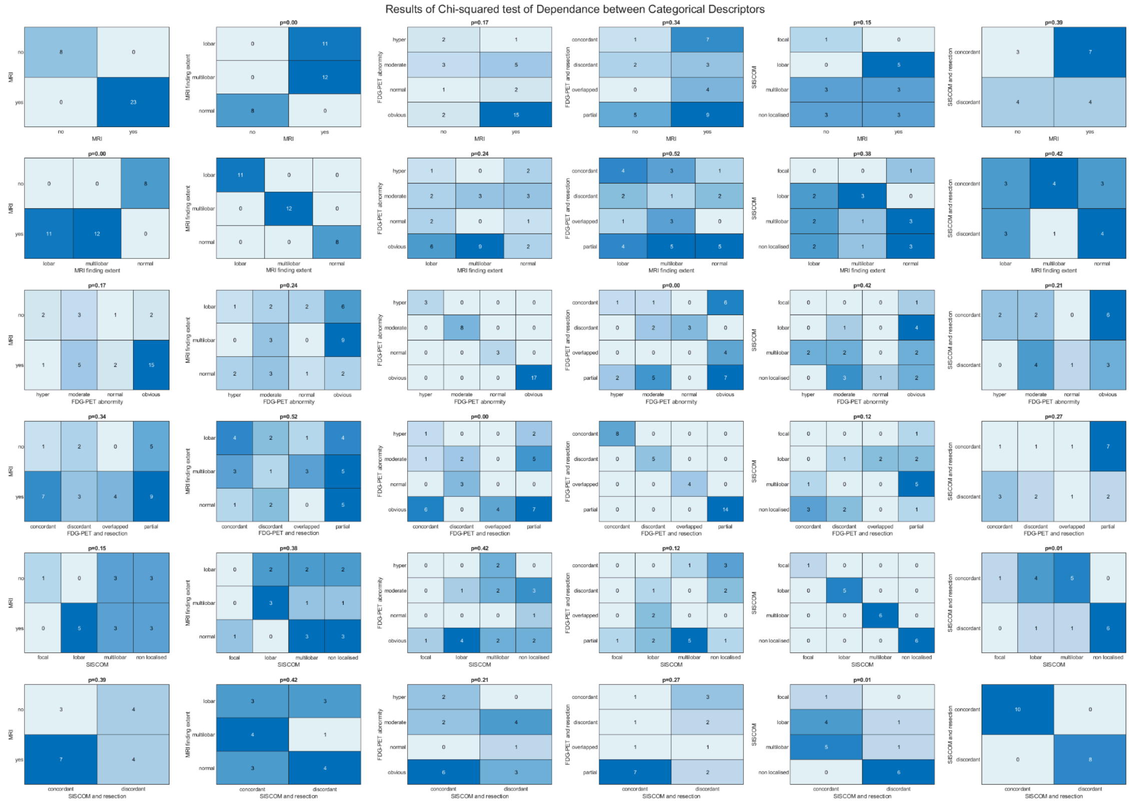


Supplementary Figure 11 shows results of Chi-squared tests between pairs of categorical descriptors. In each subplot, the data are represented by a contingency table and the test p-value is reported in the title.
